# Confidence in visual detection, familiarity and recollection judgements is preserved in schizophrenia spectrum disorder

**DOI:** 10.1101/2023.03.28.23287851

**Authors:** Martin Rouy, Michael Pereira, Pauline Saliou, Rémi Sanchez, Wassila el Mardi, Hanna Sebban, Eugénie Baqué, Childéric Dezier, Perrine Porte, Julia Micaux, Vincent de Gardelle, Pascal Mamassian, Chris J.A. Moulin, Clément Dondé, Paul Roux, Nathan Faivre

## Abstract

An effective way to quantify metacognitive abilities is to ask participants to estimate their confidence in the accuracy of their response during a cognitive task. A recent meta-analysis^1^ raised the issue that most assessments of metacognitive abilities in schizophrenia spectrum disorders may be confounded with cognitive deficits, which are known to be present in this population. Therefore, it remains unclear whether the reported metacognitive deficits are metacognitive in nature, or rather inherited from cognitive deficits. Arbitrating between these two possibilities requires equating task performance between experimental groups. Here, we aimed to characterize metacognitive performance among individuals with schizophrenia across three tasks (visual detection, familiarity, recollection) using a within-subject design, while controlling experimentally for intra-individual task performance and statistically for between-subject task performance. In line with our hypotheses, we found no metacognitive deficit for visual detection and familiarity judgements. While we expected metacognition for recollection to be specifically impaired among individuals with schizophrenia, we found evidence in favor of an absence of a deficit in that domain also. The clinical relevance of our findings is discussed in light of a hierarchical framework of metacognition.

## Introduction

Confidence abnormalities in the form of overconfidence in errors in schizophrenia spectrum disorder have been documented in multiple cognitive domains, including memory, perception, and emotion recognition^2^. Yet, the hierarchical level at which these abnormalities occur is still unclear. In line with the terminology proposed by Galvin and colleagues^3^, cognitive performance is referred to as first-order performance (i.e. how well one is able to detect or discriminate between probed stimuli), and metacognitive performance is referred to as second-order performance (i.e. how well one is able to discriminate between correct and incorrect responses). Properly quantifying metacognitive abilities requires controlling for variations of cognitive performance that are not metacognitive in nature^3–5^. This concern is of particular relevance in schizophrenia, where cognitive deficits are well documented^6,7^. In a meta-analysis we recently conducted^1^, metaperception was mostly preserved when first-order performance was controlled for. Yet, conclusions about the metamemory deficit could not be drawn in this meta-analysis since the medium to large effect size resulted from studies where memory performance was not equated between patients and healthy controls (except for one study^8^). In these conditions, metamemory deficits were likely to be confounded with memory deficits.

To compare metaperceptual and metamemory deficits in individuals with schizophrenia while controlling for perceptual and memory deficits, we developed a novel experimental paradigm including three randomly interleaved perceptual and memory tasks attempting to experimentally match first-order performance at the intra-individual level across tasks, and to statistically control for performance at the inter-individual level.

We preregistered our main predictions based on current knowledge regarding the cognitive architecture of perception and memory and their impairments in schizophrenia (see^9^ for a meta-analysis). Individuals with schizophrenia typically have preserved performance in familiarity judgements (i.e. decontextualized memory^10^) but impaired performance in recollection judgments (i.e. episodic/recollection memory necessitating multimodal integration via hippocampal activity^11^), which may be explained by impaired hippocampus recruitment^12^ and hippocampal atrophy^13^. Our main preregistered hypothesis was that metamemory was globally more impaired than metaperception, assuming that previous reports of deficits in metamemory were not only driven by deficits taking place at the first-order level. Furthermore, since familiarity can be considered a perceptual-mnemonic process storing decontextualized perceptual elements^14^, we hypothesized domain-generality between perception and familiarity processes, and expected that meta-recollection would be specifically impaired. Besides this preregistered hypothesis, we explored the links between metacognitive performance and clinical traits such as positive, negative and disorganization syndromes.

## Methods

The present design, hypotheses, and analyses were preregistered prior to data collection and analysis (https://osf.io/k4p79). Data and analysis scripts are available online (https://gitlab.com/nfaivre/metaface_scz_public).

### Participants

Following a preregistered open-ended sequential Bayes Factor design (see SI for details), we recruited 38 individuals with schizophrenia and 39 healthy control participants matched for age, sex, education level and premorbid IQ (see Table 1 for demographic and clinical information). After exclusions according to preregistered criteria (essentially due to ceiling performance, see SI for details), the analyses were conducted on a sample of 34 individuals with schizophrenia and 36 healthy controls. Two licensed psychiatrists (CD and PR) confirmed the diagnoses in the schizophrenia group according to the DSM-V criteria for schizophrenia (details about the recruitment procedure are provided in SI).

**Table 1:** Sociodemographic and clinical characteristics of individuals with schizophrenia and control participants. WAIS: Wechsler Adult Intelligence Scale (standardized scores); BCIS: Beck Cognitive Insight Scale; SSTICS: Subjective Scale To Investigate Cognition in Schizophrenia; PANSS: Positive And Negative Symptoms in Schizophrenia. p-values are not corrected for multiple comparisons. Bayes factors are based on Bayesian t-tests with a scaling factor of 0.7.

|  | Control<br>N = 36<br>(mean ± SD) | Schizophrenia<br>N = 34<br>(mean ± SD) | t-statistic | p-value | Bayes<br>Factor |
| --- | --- | --- | --- | --- | --- |
| Age, yr | 34.5 ± 14.3 | 38.3 ± 11.1 | 1.26 | 0.21 | 0.48 |
| Education Level, yr | 12.9 ± 1.3 | 12.8 ± 2.7 | -0.16 | 0.87 | 0.26 |
| Premordid IQ | 108.2 ± 6.8 | 104.9 ± 12.5 | -1.30 | 0.20 | 0.55 |
| WAIS Matrix subtest | 9.6 ± 2.9 | 8.4 ± 2.5 | -1.87 | 0.07 | 1.07 |
| Calgary Depression Scale, score | 1.5 ± 1.7 | 4.7 ± 3.9 | 4.20 | <0.001 | 209.00 |
| Illness duration, yr |  | 14.7 ± 9.3 |  |  |  |
| BCIS, composite score |  | 8.2 ± 6.2 |  |  |  |
| SSTICS, total |  | 30.1 ± 16.1 |  |  |  |
| SSTICS, working memory |  | 4.6 ± 2.7 |  |  |  |
| PANSS, positive |  | 13.2 ± 6.3 |  |  |  |
| PANSS, negative |  | 11.7 ± 9.8 |  |  |  |
| PANSS, disorganization |  | 21.7 ± 7.8 |  |  |  |
| PANSS, total |  | 48.8 ± 33.7 |  |  |  |

### Experimental design

A video description of each task is available online (https://gitlab.com/nfaivre/metaface_scz_public/-/tree/main/videos). All participants were naive to the purpose of the study, gave written informed consent in accordance with institutional guidelines and the Declaration of Helsinki, and received monetary compensation (10€ / h) except those participants under legal protection. The study was approved by the ethical committee *Sud Méditerranée* II on April the 3d 2020 (217 R01 MS1).

#### Stimuli

4000 copyright-free artificially generated faces were downloaded from the open platform https://generated.photos. Two independent observers screened the stimuli to exclude children’s faces, unrealistic faces, and faces with salient features (e.g. sunglasses, hats). The remaining 1700 male and 1700 female adult faces were converted to grayscale and equalized in contrast and luminosity (SHINE Matlab toolbox^15^). Each face was presented against a visual background noise consisting of its phase-scrambled version. The background was colorized in blue or red (balanced for luminosity) to provide a contextual cue. Size and gaze position were kept identical across all stimuli.

#### Procedure

##### Memory tasks

The familiarity and recollection tasks shared the same timeline (Figure 1). Each trial started with an encoding phase consisting of four successive face stimuli presented during 400ms each (random combination of 2 male and 2 female faces) on a blue or red background (context), with a 500 ms inter-stimulus interval. To avoid learning effects and familiarity confounds, each face was presented only once throughout the whole experiment. Following the encoding phase, the test phase consisted in presenting a fifth face on a gray background, and asking a task-specific question. In familiarity trials, the participant was asked to indicate whether the face had already been seen (80% of the trials, to obtain a uniform distribution across “stimulus strength” levels, see next paragraph) or not (20% of the trials); in recollection trials the fifth face was always a seen face (i.e. a face presented during the encoding phase), and the participant was asked whether the context of this stimulus was blue (80% of the trials) or not (20% of the trials) during the encoding phase. Participants provided their answers with a mouse click on “no” or “yes” buttons respectively displayed at the top left and top right of the screen.

**Figure 1.**
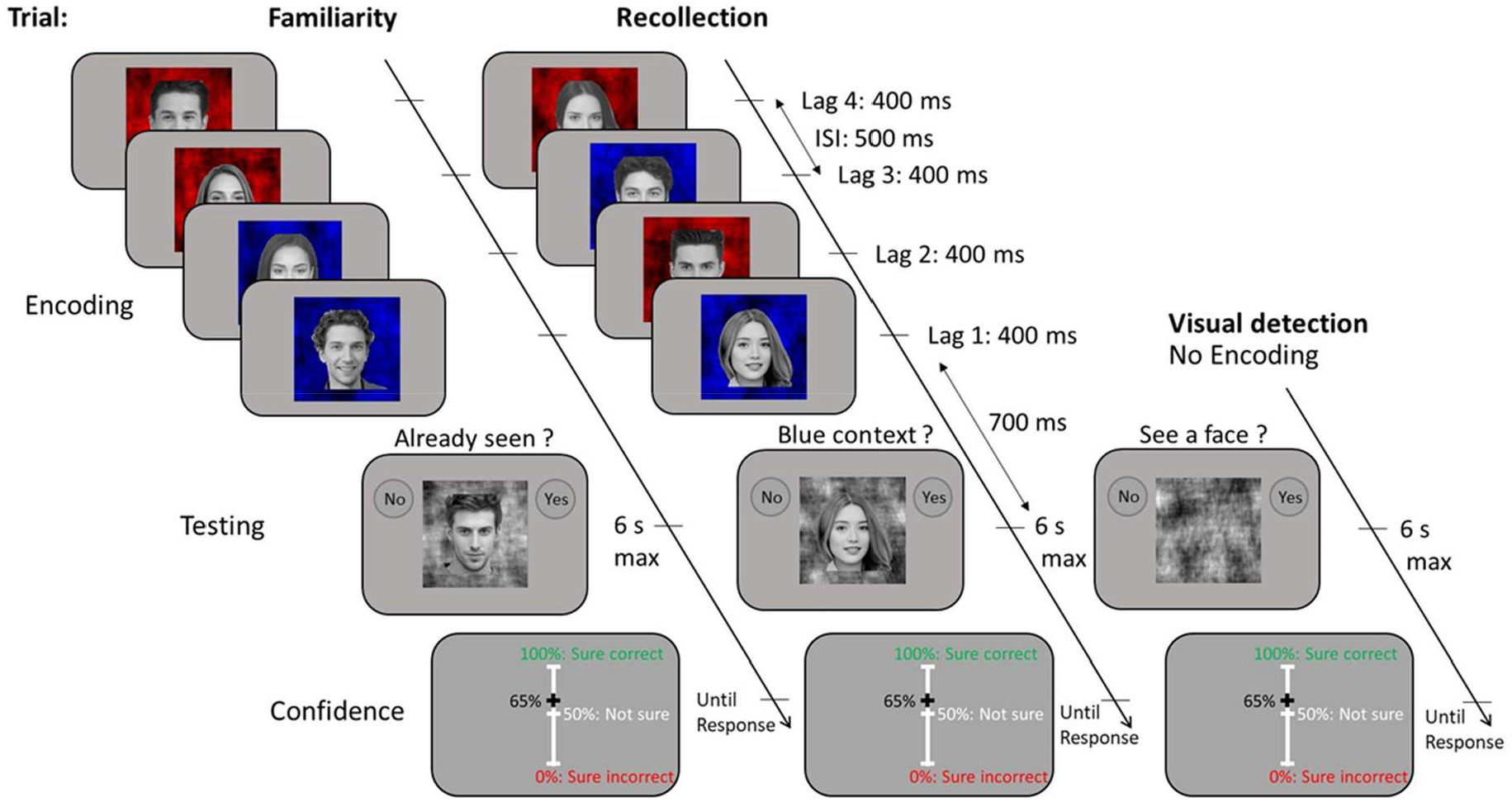
Experimental Design. Timeline of the familiarity, recollection and visual detection tasks. The timeline was identical in the familiarity and recollection task, except for the testing phase where the question was task-specific: “Already seen?” for familiarity, and “Blue context?” for recollection. No encoding took place in the visual detection task. In the present illustration, the correct answers to the familiarity, recollection and perceptual questions are respectively: “No”, “Yes”, and “No”. Lag is an ordinal variable corresponding to the temporal distance between the target stimulus and the test stimulus.

The difficulty of the familiarity and recollection tasks was manipulated by changing the serial position of the target stimulus during the encoding phase. Accordingly, there were four levels of stimulus strength - ranging from 1 to 4 -, corresponding to each of the four faces displayed sequentially within the encoding phase (Figure 1). Because this variable corresponds to the temporal distance between the target stimulus and the test stimulus, we refer to it as a “lag”. For instance, if the target face was the first face displayed during the encoding sequence, then the temporal distance between the target and the test was maximal, and the trial was categorized as “lag 4”. On the contrary, if the target was the last face of the encoding phase, the temporal distance between the target and the test was minimal, and the trial was categorized as “lag 1”. A fifth lag-level “lag 0” was used to indicate catch trials (20% of the trials): i.e. new faces in familiarity trials, faces presented in the red context in recollection trials.

##### Visual detection task

Participants had to indicate whether a face was present (80% of the trials) or not (catch trials: 20%). The face could be presented at four contrast levels, chosen to match performances obtained in the memory tasks for each of the four lags (See SI Figure S2 D). A fifth level - stimulus strength 0 - was used to tag catch trials: trials where no face was presented (20% of the trials). As for memory trials, participants provided their answer with a mouse click on “no” or “yes” buttons displayed at the top left and top right of the screen.

##### Trial exclusions

A time limit of 6 seconds was set on all trials to avoid differences in response rates between patients and controls. When the time limit was reached, an error-like sound was produced along with a visual warning in red characters asking participants to respond quicker. Proportions of non-responses were comparable between individuals with schizophrenia (mean ± SD: 2.91% ± 3.60%) and controls (mean ± SD: 2.32% ± 7.66%, BF = 0.26). These non-response trials were excluded from our analyses.

##### Confidence rating

For all three tasks, participants were asked to provide confidence judgments. After each first-order response (i.e. responses given to the familiarity, recollection and visual detection tasks) participants were asked to report their subjective confidence regarding the correctness of their decision by moving a slider with the mouse on a visual analog scale (see Figure 1) ranging from 0% (“Sure incorrect”) to 100% (“Sure correct”). The initial position of the cursor for each trial corresponded to 50% confidence (“Not sure”).

#### Structure of the experiment

This protocol aimed to match intra-individual performance across familiarity, recollection, and visual detection tasks. Participants were asked to perform two sessions of one hour each. Session 1 allowed us to measure memory performance at four difficulty levels (according to the variable “lag”, see *Memory Tasks)*. We then matched perceptual performance to memory performance by determining four adequate contrast levels for the visual detection task for each participant (see SI for details). Thus, session 1 provided four levels of stimulus strength, i.e. 4 memory lags and 4 visual contrast levels, corresponding to matched performance for each participant. Based on these individual parameters, session 2 contained 10 blocks of 30 randomly interleaved trials (familiarity, recollection and visual detection task), totalizing 300 trials (100 trials per task), each followed by a confidence rating task. Task order and stimulus strength were randomized, so participants could not predict which task they were going to perform on each trial.

Importantly, this paradigm was designed to match first-order performance between tasks, which is convenient to compare metacognitive deficits across tasks. Although we also attempted to match first-order performance between groups, pilot experiments revealed this was not possible using adaptive staircases. Therefore, differences in task performance between groups were accounted for at the statistical level using the confidence efficiency metric^16^, taking advantage of our design with different levels of difficulty.

### Statistical analyses

Analyses were performed with R^17^, using notably the brms^18^, BayesFactor^19^, ggplot2^20^ and lme4^21^ packages. Confidence efficiency scores were computed with Matlab (Mathworks, 2017a).

#### Socio-demographic and neuropsychological characterization

The groups’ socio-demographic (age, sex, education), neuropsychological (National Adult Reading Test measuring patients’ premorbid IQ^22^, matrix reasoning subtest from the Wechsler Adult Intelligence Scale version IV^23^ and mood (Calgary Depression Scale^24^) characteristics were compared using the Student t test or χ^2^ test when appropriate. Patients were characterized in terms of cognitive insight (using the self-reported Beck Cognitive Insight Scale^25^), schizophrenia symptomatology (using the clinician-evaluated Positive And Negative Syndrome Scale^26^, with factorial scores^27^) and subjective evaluation of cognitive functioning (using the self-reported Subjective Scale To Investigate Cognition in Schizophrenia^28^). As additional analyses, we explored whether metacognitive performance was correlated with demographic characteristics and clinical scores.

#### Metacognitive performance

We quantified metacognitive sensitivity with population-level estimates of confidence efficiency^16^. This model accounts for potential differences in first-order performance and relies on an explicit generative model of confidence. We also quantified metacognitive sensitivity with a measure of the strength of the relationship between first-order accuracy and confidence (via individual regression slopes), obtained from Bayesian mixed-effects logistic regressions. Importantly, this second model did not take first-order performance into account. Thus, comparing the two measures of metacognitive sensitivity, we assessed the importance of controlling for first-order performance.

##### Bayesian mixed-effects logistic regressions

We conducted two Bayesian mixed-effects logistic regressions on first-order accuracy (binary categorical variable) as a function of standardized confidence (continuous variable): one model (1a, see below) for *hit* vs *miss* responses (i.e. stimulus strength [1:4]), and one model (1b) for *false alarms* vs. *correct rejections* (i.e. stimulus strength = 0). We analyzed trials with 0 stimulus strength separately (i.e., 0 versus [1:4]) assuming that stimulus strength 0 involved different processes (e.g. detecting a new stimulus may not be based on the same information as detecting an old stimulus. This was corroborated by pilot experiments showing that task-performance at stimulus strength 0 was hardly extrapolated from stimulus strength > 0). Model 1a included group (binary categorical variable: controls vs patients), stimulus strength (ordinal variable with 4 levels: 1 to 4), task (categorical variable: visual detection, familiarity, recollection) as fixed effects, and a full random effect structure (see SI for priors’ specifications). Model 1b included the same variables except that stimulus strength was fixed to 0. Results were interpreted on the basis of the Bayes factor (BF) according to Wagenmakers and colleagues^29^. The BF is the ratio of the marginal likelihoods of each hypothesis, therefore BF > 3 indicates evidence toward H1 (existence of a difference between conditions) and BF < 1/3 indicates evidence toward H0 (absence of difference between conditions). Effects were further characterized by the summary statistics of the posterior distribution (mean and 95 % credible interval, CrI).

Formulae:

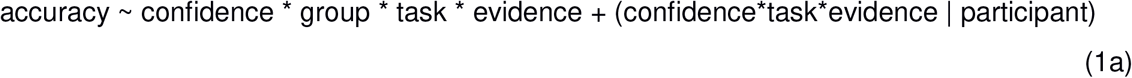

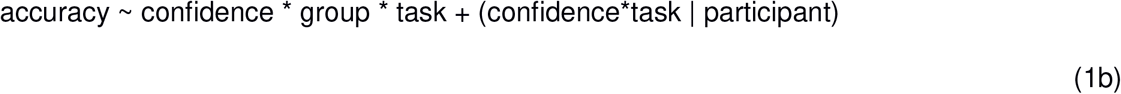

##### Confidence efficiency

As preregistered, we assessed metacognitive performance while accounting for first-order performance and task difficulty with a recently developed metacognitive index called “confidence efficiency”^16^, here adapted to confidence ratings. This index is based on a generative model of confidence judgments, based on Signal Detection Theory, where observers’ confidence judgments are not only subject to metacognitive noise but may also incorporate additional information from the stimulus. Interestingly for us, this method enables the simultaneous modeling of confidence responses across different levels of task difficulty, unlike other methods such as M-ratio^4,5^.

We estimated confidence efficiency by collapsing all participants into one global population, after normalizing for variations in task performance across individuals, and we quantified its dispersion using a bootstrapping procedure. Namely, we computed 1000 confidence efficiency estimates based on a random resampling of our pool of participants (with replacement), resulting in one estimation distribution per task and group.

Our predictions regarding metacognitive performance (i.e., confidence efficiency and slopes of mixed-effects logistic regressions) were as follows: 1) A metamemory deficit for individuals with schizophrenia compared to healthy controls. 2) A significant interaction effect between group and task reflecting a larger deficit in recollection metamemory among individuals with schizophrenia compared to other tasks, whereas healthy controls show no differences in metacognitive performances across tasks.

We also expected intra-individual first-order performances to be matched (assessed with model 1a), as reflected by equivalent accuracy across the three tasks among patients and healthy controls. Since we did not experimentally adapt task performance between groups, we expected lower task performances among patients compared to controls.

## Results

### Clinical and neuropsychological variables

Groups were balanced for sex (χ^2^ = 0.25, p = 0.62) and comparable for age, education level, premorbid IQ, and scores on the WAIS matrix subtest (Table 1). However, individuals with schizophrenia had higher depression scores (mean ± SD: 4.7 ± 3.9) than healthy controls (mean ± SD: 1.5 ± 1.7, t = 4.20, p < 0.001, BF = 209). Descriptive statistics regarding false alarms, hits and confidence are described in Table 2 and show that in both groups, participants were performing all tasks correctly (i.e., better than chance).

**Table 2:** Experimental characteristics of individuals with schizophrenia and healthy control participants. p-values are not corrected for multiple comparisons. Bayes factors are based on Bayesian t-tests with a scaling factor of 0.7.

|  | Task | Control<br>N = 36<br>(mean ± SD) | Schizophrenia<br>N = 34<br>(mean ± SD) | t-statistic | p-value | Bayes<br>Factor |
| --- | --- | --- | --- | --- | --- | --- |
| % False alarms | Visual detection | 20.0 ± 20.7 | 16.9 ± 24.5 | -0.57 | 0.57 | 0.28 |
|  | Familiarity | 17.1 ± 14.1 | 24.6 ± 21.1 | 1.74 | 0.09 | 0.92 |
|  | Recollection | 27.9 ± 17 | 50.6 ± 23.3 | 4.63 | 0.00 | 1,211.87 |
| % Hits | Visual detection | 80.2 ± 13 | 70.6 ± 18.4 | -2.50 | 0.02 | 3.53 |
|  | Familiarity | 81.3 ± 10.4 | 72.8 ± 20.1 | -2.20 | 0.03 | 2.02 |
|  | Recollection | 78.5 ± 15.9 | 72.3 ± 14.9 | -1.68 | 0.10 | 0.82 |
| % Confidence in errors | Visual detection | 82.8 ± 11.5 | 85.6 ± 11.4 | 1.02 | 0.31 | 0.39 |
|  | Familiarity | 73.1 ± 12 | 72.7 ± 11.2 | -0.13 | 0.89 | 0.25 |
|  | Recollection | 73.2 ± 12.1 | 70.5 ± 11 | -0.97 | 0.33 | 0.37 |
| % Confidence in correct responses | Visual detection | 95.0 ± 5.3 | 91.6 ± 9.7 | -1.82 | 0.07 | 1.06 |
|  | Familiarity | 89.2 ± 6.8 | 84.6 ± 11 | -2.07 | 0.04 | 1.57 |
|  | Recollection | 87.4 ± 10 | 80.9 ± 10.9 | -2.58 | 0.01 | 4.04 |

### First-order performance

Model 1a revealed that patients had lower performance than healthy controls in the visual detection, familiarity and recollection tasks, and these first-order deficits were similar across tasks (i.e. no first-order interactions, see Table 3, Figure 2A).

**Table 3:** First-order deficits across tasks. We report posterior distributions’ summary statistics (mean and 95% Credible interval) along with Bayes factors.

|  |  | Estimate [95% CrI] | Bayes Factor |
| --- | --- | --- | --- |
| First-order deficits | Schizophrenia-Control (Visual detection) | -0.82 [-1.43, -0.21] | 8.40 |
|  | Schizophrenia-Control (Familiarity) | -0.81 [-1.49, -0.13] | 3.07 |
|  | Schizophrenia-Control (Recollection) | -0.62 [-1.27, 0.02] | 1.23 |
| First-order interactions | group x task (Familiarity - Visual detection) | 0.01 [-0.57, 0.62] | 0.31 |
|  | group x task (Recollection - Visual detection) | 0.2 [-0.38, 0.78] | 0.38 |
|  | group x task (Recollection - Familiarity) | 0.19 [-0.41, 0.81] | 0.27 |
| Intra-individual performance-matching | Familiarity - Visual detection (Control) | 0.13 [-0.31, 0.56] | 0.26 |
|  | Recollection - Visual detection (Control) | -0.2 [-0.62, 0.21] | 0.32 |
|  | Recollection - Familiarity (Control) | -0.33 [-0.78, 0.12] | 0.50 |
|  | Familiarity - Visual detection (Schizophrenia) | 0.14 [-0.31, 0.58] | 0.20 |
|  | Recollection - Visual detection (Schizophrenia) | 0 [-0.44, 0.43] | 0.16 |
|  | Recollection - Familiarity (Schizophrenia) | -0.14 [-0.59, 0.31] | 0.14 |

**Figure 2:**
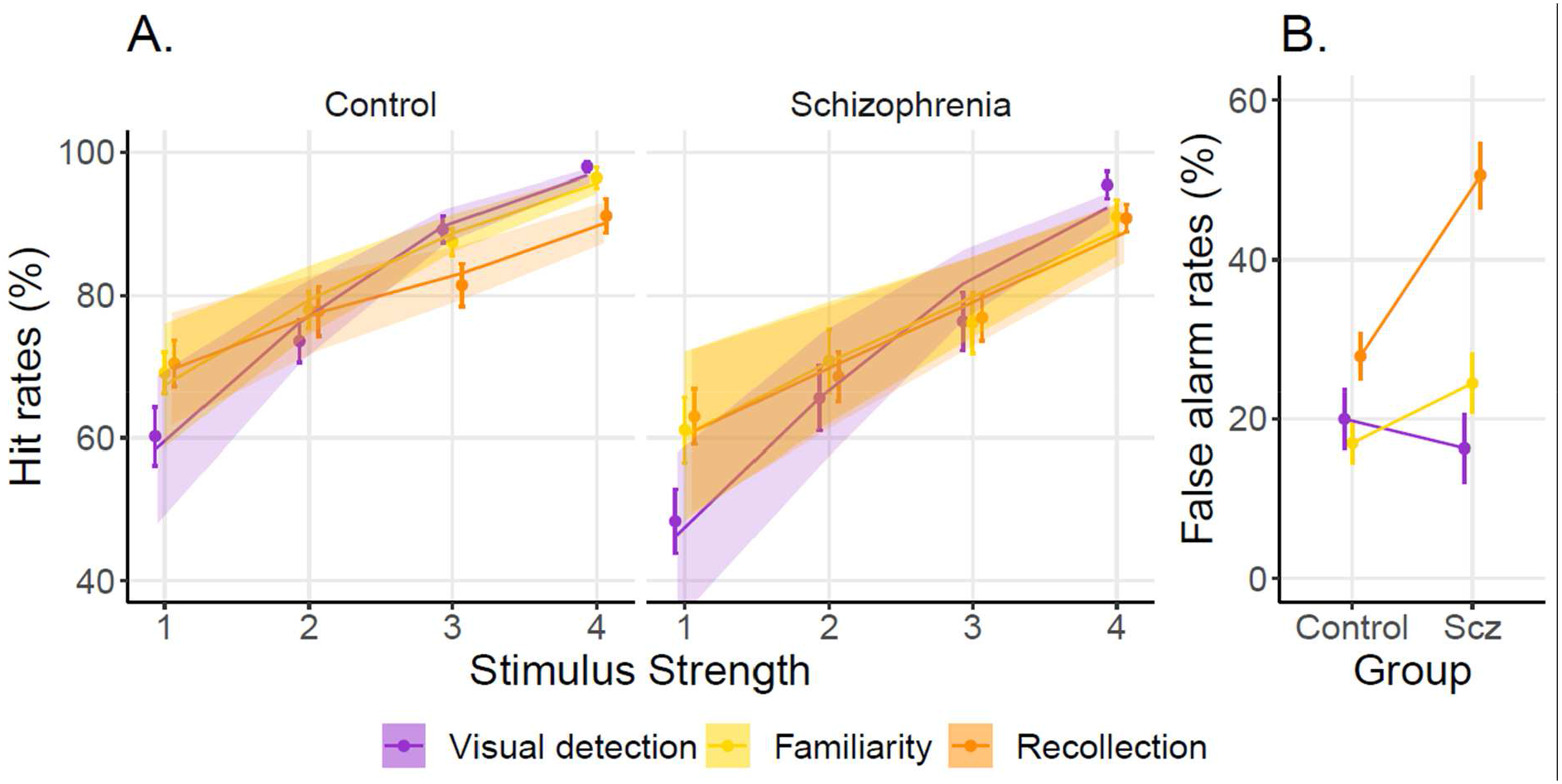
A. Hit rates (i.e., rates of “yes” responses following stimuli with stimulus strength > 0) across stimulus strengths in the visual detection (purple), familiarity (yellow), and recollection tasks (orange). Points and error bars indicate average accuracy and standard error of the mean, respectively; solid lines and shaded areas represent model fit mean and 95% confidence interval, respectively. B. False-alarm rates (i.e., rates of “yes” responses following stimuli with 0 stimulus strength) across groups. Points and error bars indicate average accuracy and standard error of the mean, respectively. Same color description as panel A.

Differences in performance were expected as task performance was not experimentally controlled between groups. However, our procedure was designed to match intra-individual performance across tasks. Accordingly, pairwise first-order task performances were similar among patients and among control participants (Table 3). This confirms that our procedure globally matched intra-individual performance across tasks, although it did not match intra-individual performance for each stimulus strength (see Table S1).

Patients and controls were sensitive to task manipulation of stimulus strength as indicated by a strong effect of stimulus strength in all tasks (See Table S1 and Figure 2A).

#### False alarms

Compared to healthy controls, patients had a similar false alarm rate in both the visual detection task (i.e., reporting seeing a face when none was presented: -0.22 [-1.02, 0.57], BF = 0.45), and in the familiarity task (i.e., reporting having seen the test face during the encoding phase when presented with a new face: 0.78 [-0.07, 1.64], BF = 2.21) but they committed significantly more false alarms in the recollection task (i.e., reporting having seen the test face in a given context during the encoding phase when presented in another context: 1.54 [0.61, 2.46], BF = 96.7) (Figure 2B).

### Second-order performance

#### Confidence

Confidence levels were similar between patients and controls, except for the recollection task where patients were underconfident in correct responses (Table 1, confidence mean ± SD: 80.9 ± 10.9) compared to controls (confidence mean ± SD: 87.4 ± 10.0, t = -2.58, p < 0.05, BF = 4.04).

##### Metacognitive sensitivity

When quantifying metacognitive sensitivity as the slope between accuracy and confidence in mixed-effects logistic regressions (model 1a), individuals with schizophrenia were not found to underperform compared to healthy controls (Figure 3A). Although qualitatively, the results could suggest a metacognitive deficit in the visual detection task, the evidence was statistically inconclusive (−0.41 [-0.84, 0.01], BF = 1.33). By contrast, we obtained moderate evidence in favor of an absence of a deficit both in meta-familiarity (−0.24 [-0.59, 0.12], BF = 0.32), and meta-recollection (−0.13 [-0.51, 0.27], BF = 0.17). Moreover, there was no difference of deficit between tasks (Familiarity - Recollection: 0.11 [- 0.33, 0.56], BF = 0.18); Familiarity - Perception: 0.17 [- 0.26, 0.62], BF = 0.28; Recollection - Perception: 0.28 [- 0.18, 0.75], BF = 0.47). As discussed above, metacognitive sensitivity can be contaminated by differences in terms of first-order performance, which was only partially controlled in our paradigm. To estimate metacognitive performance independently of first-order performance, we turned to another metric called the confidence efficiency.

**Figure 3:**
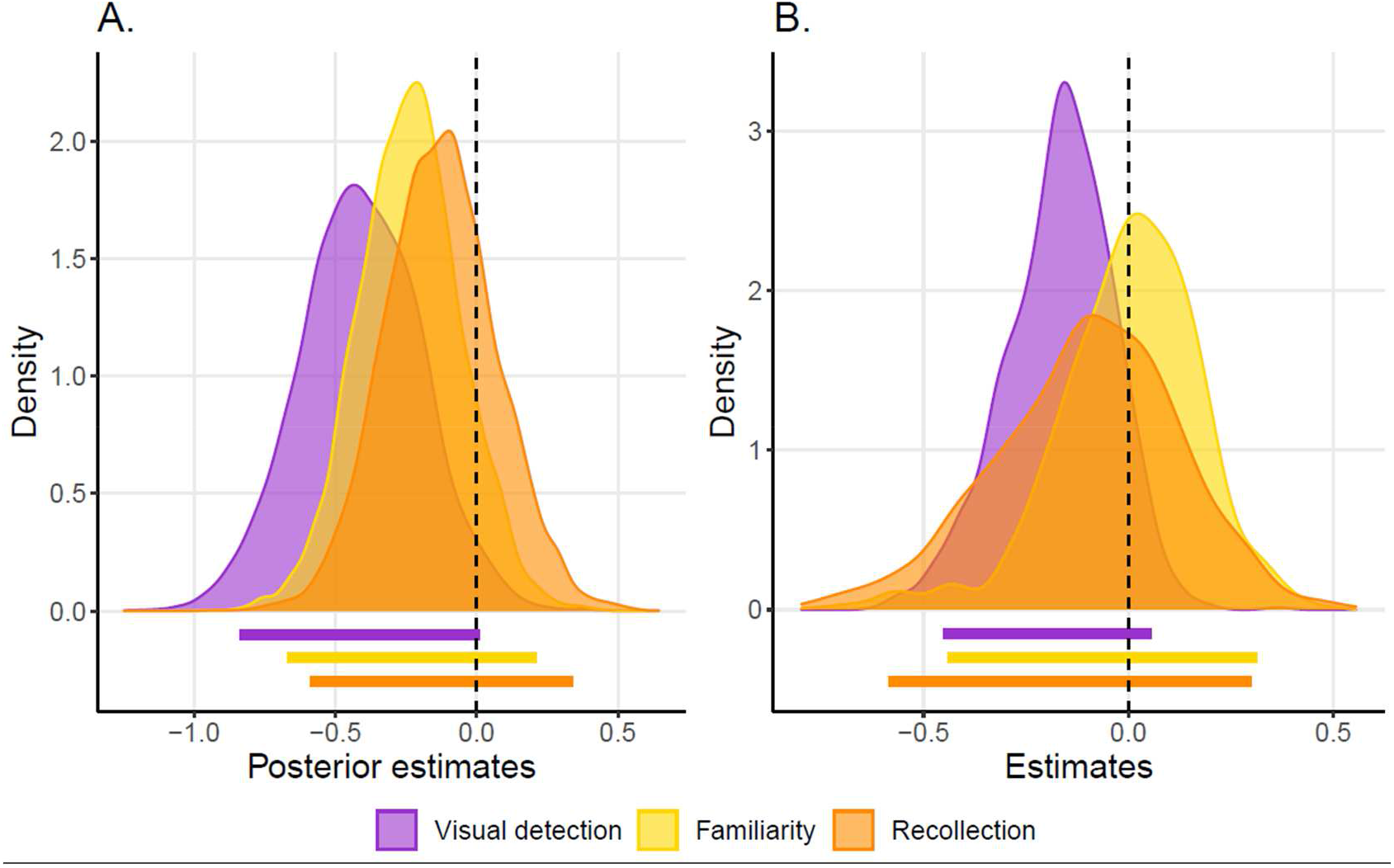
A. Bayesian posterior distributions of differences of regression slope estimates between patients and controls (i.e. distributions of metacognitive deficits estimations): meta-perceptual difference (purple), meta-familiarity difference (yellow), meta-recollection difference (orange). Vertical dashed line (estimate = 0) represents no difference between patients and controls. Horizontal colored bars indicate 95% credible intervals. B. Distributions of differences in confidence efficiency estimates between patients and controls. Horizontal colored bars indicate 95% confidence intervals. Same color description as panel A.

When quantifying metacognitive performance using the confidence efficiency measure of metacognition - which controls for first-order deficits - individuals with schizophrenia had similar confidence efficiency in the detection (−0.17 [-0.45, 0.06]), familiarity (−0.00 [-0.44, 0.31]) and recollection tasks (−0.10 [-0.58, 0.30]) (Figure 3B). Within each group, metacognitive performance was comparable across tasks (Controls: Visual detection - Familiarity: -0.11[-0.40, 0.20], Visual detection - Recollection: -0.18[-0.51, 0.17], Recollection - Familiarity 0.07[-0.26, 0.41]; Patients: Visual detection - Familiarity: -0.28[-0.55, 0.14], Visual detection - Recollection: -0.24[-0.57, 0.19], Recollection - Familiarity -0.03[-0.50, 0.42]).

Contrary to our predictions about domain-generality, metacognitive performance as measured with mixed-effects logistic regressions did correlate across tasks, neither did we find correlations with clinical traits such as positive, negative and disorganization syndromes and cognitive insight (see SI, Figure S7, 8).

## Discussion

The present study aimed at characterizing metamemory and metaperception in people with schizophrenia while controlling for first-order deficits. In particular, we assessed metacognition in visual detection, familiarity, and recollection tasks. We hypothesized that people with schizophrenia would be specifically impaired in the meta-memory domain. At the first-order level, we found that people with schizophrenia had lower first-order performance in the three tasks compared to healthy controls, which confirms the importance of accounting for first-order deficits to quantify second-order processes specifically. When doing so, contrary to our hypothesis we found that metacognitive sensitivity was preserved among individuals with schizophrenia in the three tasks. In what follows, we discuss technical and conceptual aspects of our paradigm that should be considered to interpret this result, and then examine its clinical and theoretical significance.

A key contribution of this study is our attempt to match first-order performance between tasks for each participant using adaptive procedures, and between groups of participants using a generative model of confidence. We note that our adaptive procedure to match performance between tasks was successful when considering average performance, but not when considering task performance across levels of stimulus strength. In other words, we equated the overall performance but not the slopes between tasks in Figure 2A (see SI for details). A plausible explanation for this is a contextual effect. In session 1, blocks of visual detection trials were separated from blocks of memory trials, whereas in session 2 the three tasks were interleaved within each block of trials. Thus, the visual detection psychometric curve (SI, Figure S2c) from which we determined four visual contrast levels was obtained from a sequence of low-contrast perceptual stimuli (3 × 80 stimuli in a row), whereas during session 2 these low-contrast visual stimuli were interleaved with high-contrasted memory stimuli. This contextual effect might have resulted in a rightward shift (See Figure S3) of the visual detection psychometric curve, leading to underperformance in both groups in the visual detection task compared to the familiarity and recollection tasks.

Regarding between-groups task performance matching, early pilot versions of the present protocol aimed at equating memory performance between participants using adaptive staircases that manipulated either the number of encoding items, or the lag variable, but these attempts were not successful (no convergence). Instead, we accounted for differences in task-performance between groups by relying on measures of confidence efficiency from a recent generative model of confidence^16^, which enables the estimation of metacognitive abilities in factorial designs. Although this framework is recent and has not been fully benchmarked yet, we note that we found qualitatively similar results using a Bayesian logistic mixed-effects regression, which does not consider possible cognitive deficits but has the advantage of providing hierarchical estimates of metacognitive sensitivity, dealing with unbalanced data, and considering prior knowledge to compute Bayes factors. In contradiction to existing literature, both frameworks revealed no evidence for a metacognitive deficit in any of the three tasks. In fact, we found evidence for an absence of metacognitive deficit in memory tasks, and only inconclusive evidence in the perceptual domain. The absence of metacognitive deficit in schizophrenia was corroborated by an absence of difference regarding confidence biases. Indeed, contrary to several studies which did not control for first-order performance^30–32^, we found no overconfidence in errors nor underconfidence in correct responses. One possibility is that the confidence biases previously reported in schizophrenia also stem from first-order deficits differences. Furthermore, contrary to previous behavioral results showing a positive link between false alarms and positive symptoms or proneness to hallucinations^33–35^, our sample of patients had comparable rates of false alarms compared to healthy controls in the visual detection task. They committed more false-alarms in the memory tasks, interpreted as false recognitions, but no relationships were found between rates of false alarms and PANSS positive score (see SI).

At a conceptual level, the framing of our memory tasks in terms of familiarity and recollection processes may be questionable. Indeed, although our recognition memory tasks shared some features with usual familiarity and recollection tasks (in particular the testing questions which are respectively context-independent and context-dependent), there was no delay between encoding and testing phases, as we manipulated task difficulty with a variable lag. Therefore, one may consider our tasks to reflect working memory, which is also known to involve familiarity and recollection processes^36^. To our knowledge, no study assessed metacognition related to short-term memory in schizophrenia. At first sight, our results seem to be in contradiction with the study by Berna et and colleagues^8^, which reported impaired metamemory in schizophrenia in a long-term (autobiographical) memory task, while controlling statistically for first-order performance. Yet, if our results are construed as evidence for preserved “short-term” metamemory in schizophrenia, the contradiction might be only apparent. A full taxonomy of metamemory processes is beyond the scope of the present study, and developing new paradigms to assess metacognitive performance in distinct subdomains of memory while controlling for first-order performance is one of the numerous challenges the metacognitive field is facing^37^.

With these technical and conceptual considerations in mind, we can contextualize our findings and assess their clinical relevance. Our protocols focus on “in-the-moment” metacognition^38^, i.e. confidence in trial-by-trial decisions, also known as “local” metacognition as opposed to more “global” evaluations^39–41^. Metacognitive evaluations have been construed as hierarchically organized, where aggregated local judgments give rise to global self-beliefs about one’s performance within a cognitive task or domain^42^. Interestingly, it has been shown that global metacognitive evaluations can be altered independently from the local monitoring processes^43^. Yet, as recently discussed^44^, both local and global measures of metacognition may give an incomplete picture of metacognitive abilities from a clinical perspective. This concern is corroborated by the fact that our metacognitive measures are not correlated with several clinical dimensions of interest for schizophrenia (symptoms, cognitive insight, self-reported cognitive functioning see SI). Only perceptual reasoning assessed with WAIS matrix subtest scores were positively correlated with metacognitive performance, as reported previously^45^. The need for paradigms that do justice to the breadth of the metacognition construct, i.e. including more cognitive domains, larger timescales, and theory of mind is now becoming acknowledged by the field.

## Data Availability

All data produced are available online at https://gitlab.com/nfaivre/metaface_scz_public

https://osf.io/k4p79

## Acknowledgments

NF has received funding from the European Research Council (ERC) under the European Union’s Horizon 2020 research and innovation programme (Grant agreement No. 803122).

## References

1. Rouy M, Saliou P, Nalborczyk L, Pereira M, Roux P, Faivre N. Systematic review and meta-analysis of metacognitive abilities in individuals with schizophrenia spectrum disorders. Neurosci Biobehav Rev. 2021;126:329–337. doi:10.1016/j.neubiorev.2021.03.017

2. Hoven M, Lebreton M, Engelmann JB, Denys D, Luigjes J, van Holst RJ. Abnormalities of confidence in psychiatry: an overview and future perspectives. Transl Psychiatry. 2019;9(1):268. doi:10.1038/s41398-019-0602-7

3. Galvin SJ, Podd JV, Drga V, Whitmore J. Type 2 tasks in the theory of signal detectability: Discrimination between correct and incorrect decisions. Psychon Bull Rev. 2003;10(4):843–876. doi:10.3758/BF03196546

4. Fleming SM, Lau HC. How to measure metacognition. Front Hum Neurosci. 2014;8. doi:10.3389/fnhum.2014.00443

5. Maniscalco B, Lau H. A signal detection theoretic approach for estimating metacognitive sensitivity from confidence ratings. Conscious Cogn. 2012;21(1):422–430. doi:10.1016/j.concog.2011.09.021

6. Gopal YV, Variend H. First-episode schizophrenia: review of cognitive deficits and cognitive remediation. Adv Psychiatr Treat. 2005;11(1):38–44. doi:10.1192/apt.11.1.38

7. Schaefer J, Giangrande E, Weinberger DR, Dickinson D. The global cognitive impairment in schizophrenia: Consistent over decades and around the world. Schizophr Res. 2013;150(1):42–50. doi:10.1016/j.schres.2013.07.009

8. Berna F, Zou F, Danion JM, Kwok SC. Overconfidence in false autobiographical memories in patients with schizophrenia. Psychiatry Res. 2019;279:374–375. doi:10.1016/j.psychres.2018.12.063

9. Libby LA, Yonelinas AP, Ranganath C, Ragland JD. Recollection and Familiarity in Schizophrenia: A Quantitative Review. Biol Psychiatry. 2013;73(10):944–950. doi:10.1016/j.biopsych.2012.10.027

10. Mishkin M, Suzuki WA, Gadian DG, Vargha–Khadem F. Hierarchical organization of cognitive memory. Burgess N, Oapos;Keefe J, eds. Philos Trans R Soc Lond B Biol Sci. 1997;352(1360):1461–1467. doi:10.1098/rstb.1997.0132

11. Moulin CJA, Souchay C, Morris RG. The cognitive neuropsychology of recollection. Cortex. 2013;49(6):1445–1451. doi:10.1016/j.cortex.2013.04.006

12. Heckers S, Rauch S, Goff D, et al. Impaired recruitment of the hippocampus during conscious recollection in schizophrenia. Nat Neurosci. 1998;1(4):318–323. doi:10.1038/1137

13. van Erp TGM, Hibar DP, Rasmussen JM, et al. Subcortical brain volume abnormalities in 2028 individuals with schizophrenia and 2540 healthy controls via the ENIGMA consortium. Mol Psychiatry. 2016;21(4):547–553. doi:10.1038/mp.2015.63

14. Barbeau EJ, Taylor MJ, Regis J, Marquis P, Chauvel P, Liegeois-Chauvel C. Spatio temporal Dynamics of Face Recognition. Cereb Cortex. 2008;18(5):997–1009. doi:10.1093/cercor/bhm140

15. Willenbockel V, Sadr J, Fiset D, Horne GO, Gosselin F, Tanaka JW. Controlling low-level image properties: The SHINE toolbox. Behav Res Methods. 2010;42(3):671–684. doi:10.3758/BRM.42.3.671

16. Mamassian P, de Gardelle V. Modeling perceptual confidence and the confidence forced-choice paradigm. Psychol Rev. Published online July 29, 2021. doi:10.1037/rev0000312

17. R Core Team. R: A Language and Environment for Statistical Computing. https://www.R-project.org/

18. Bürkner PC. brms : An R Package for Bayesian Multilevel Models Using Stan. J Stat Softw. 2017;80(1). doi:10.18637/jss.v080.i01

19. Morey RD, Rouder JN. BayesFactor: Computation of Bayes Factors for Common Designs. Published online 2018. https://CRAN.R-project.org/package=BayesFactor

20. Wickham H. ggplot2: Elegant Graphics for Data Analysis. Published online 2016. https://ggplot2.tidyverse.org

21. Bates D, Mächler M, Bolker B, Walker S. Fitting Linear Mixed-Effects Models using lme4. Published online 2014. doi:10.48550/ARXIV.1406.5823

22. Mackinnon A, Mulligan R. Estimation de l’intelligence prémorbide chez les francophones. L’Encéphale. 2005;31(1):31–43. doi:10.1016/S0013-7006(05)82370-X

23. Wechsler D. Wechsler Adult Intelligence Scale--Fourth Edition. Published online November 12, 2012. doi:10.1037/t15169-000

24. Addington D, Addington J, Maticka-tyndale E. Assessing Depression in Schizophrenia: The Calgary Depression Scale. Br J Psychiatry. 1993;163(S22):39–44. doi:10.1192/S0007125000292581

25. Beck A. A new instrument for measuring insight: the Beck Cognitive Insight Scale. Schizophr Res. 2004;68(2-3):319–329. doi:10.1016/S0920-9964(03)00189-0

26. Kay SR, Fiszbein A, Opler LA. The Positive and Negative Syndrome Scale (PANSS) for Schizophrenia. Schizophr Bull. 1987;13(2):261–276. doi:10.1093/schbul/13.2.261

27. Vandergaag M, Hoffman T, Remijsen M, et al. The five-factor model of the Positive and Negative Syndrome Scale II: A ten-fold cross-validation of a revised model. Schizophr Res. 2006;85(1-3):280–287. doi:10.1016/j.schres.2006.03.021

28. Stip E, Caron J, Renaud S, Pampoulova T, Lecomte Y. Exploring cognitive complaints in schizophrenia: the subjective scale to investigate cognition in schizophrenia. Compr Psychiatry. 2003;44(4):331–340. doi:10.1016/S0010-440X(03)00086-5

29. Wagenmakers EJ, Marsman M, Jamil T, et al. Bayesian inference for psychology. Part I: Theoretical advantages and practical ramifications. Psychon Bull Rev. 2018;25(1):35–57. doi:10.3758/s13423-017-1343-3

30. Eifler S, Rausch F, Schirmbeck F, et al. Metamemory in schizophrenia: retrospective confidence ratings interact with neurocognitive deficits. Psychiatry Res. 2015;225(3):596–603. doi:10.1016/j.psychres.2014.11.040

31. Garcia CP, Sacks SA, Weisman de Mamani AG. Neurocognition and Cognitive Biases in Schizophrenia. J Nerv Ment Dis. 2012;200(8):724–727. doi:10.1097/NMD.0b013e3182614264

32. Eisenacher S, Zink M. The Importance of Metamemory Functioning to the Pathogenesis of Psychosis. Front Psychol. 2017;8:304. doi:10.3389/fpsyg.2017.00304

33. Bentall RP, Slade PD. Reality testing and auditory hallucinations: A signal detection analysis. Br J Clin Psychol. 1985;24(3):159–169. doi:10.1111/j.2044-8260.1985.tb01331.x

34. Moseley P, Fernyhough C, Ellison A. The role of the superior temporal lobe in auditory false perceptions: A transcranial direct current stimulation study. Neuropsychologia. 2014;62:202–208. doi:10.1016/j.neuropsychologia.2014.07.032

35. Powers AR, Mathys C, Corlett PR. Pavlovian conditioning–induced hallucinations result from overweighting of perceptual priors. Science. 2017;357(6351):596–600. doi:10.1126/science.aan3458

36. Oberauer K. Binding and Inhibition in Working Memory: Individual and Age Differences in Short-Term Recognition. J Exp Psychol Gen. 2005;134(3):368–387. doi:10.1037/0096-3445.134.3.368

37. Rahnev D, Balsdon T, Charles L, et al. Consensus Goals in the Field of Visual Metacognition. PsyArXiv; 2021. doi:10.31234/osf.io/z8v5x

38. Palmer-Cooper EC, Wright AC, McGuire N, et al. Metacognition and psychosis-spectrum experiences: A study of objective and subjective measures. Schizophr Res. Published online January 2023:S0920996422004613. doi:10.1016/j.schres.2022.12.014

39. Lee ALF, de Gardelle V, Mamassian P. Global visual confidence. Psychon Bull Rev. 2021;28(4):1233–1242. doi:10.3758/s13423-020-01869-7

40. Rouault M, Fleming SM. Formation of global self-beliefs in the human brain. Proc Natl Acad Sci. 2020;117(44):27268–27276. doi:10.1073/pnas.2003094117

41. Cavalan Q, Vergnaud JC, de Gardelle V. From local to global estimations of confidence in perceptual decisions. Forthcoming in JEP General.

42. Seow TXF, Rouault M, Gillan CM, Fleming SM. Reply to: Metacognition, Adaptation, and Mental Health. Biol Psychiatry. 2022;91(8):e33–e34. doi:10.1016/j.biopsych.2021.11.005

43. Bhome R, McWilliams A, Price G, et al. Metacognition in functional cognitive disorder. Brain Commun. 2022;4(2):fcac041. doi:10.1093/braincomms/fcac041

44. Schnakenberg Martin AM, Lysaker PH. Metacognition, Adaptation, and Mental Health. Biol Psychiatry. 2022;91(8):e31–e32. doi:10.1016/j.biopsych.2021.09.028

45. Faivre N, Roger M, Pereira M, et al. Confidence in visual motion discrimination is preserved in individuals with schizophrenia. J Psychiatry Neurosci. 2021;46(1):E65–E73. doi:10.1503/jpn.200022

