## Supplementary information for "Confidence in visual detection, familiarity and recollection judgements is preserved in schizophrenia spectrum disorder"

### Methods

#### Participants

##### Optional bayesian stopping rule:

As per our preregistered plan, we sought to include 50 patients and 50 healthy controls, or to stop the recruitment whenever moderate evidence for the presence (BF > 3) or absence (< 0.33) of a specific metacognitive impairment among individuals with schizophrenia indicated by an interaction between group and task-domain on metaperformance. It turned out that our preregistered evidence thresholds were already exceeded when we fully inspected the data for the first time (Figure S1).

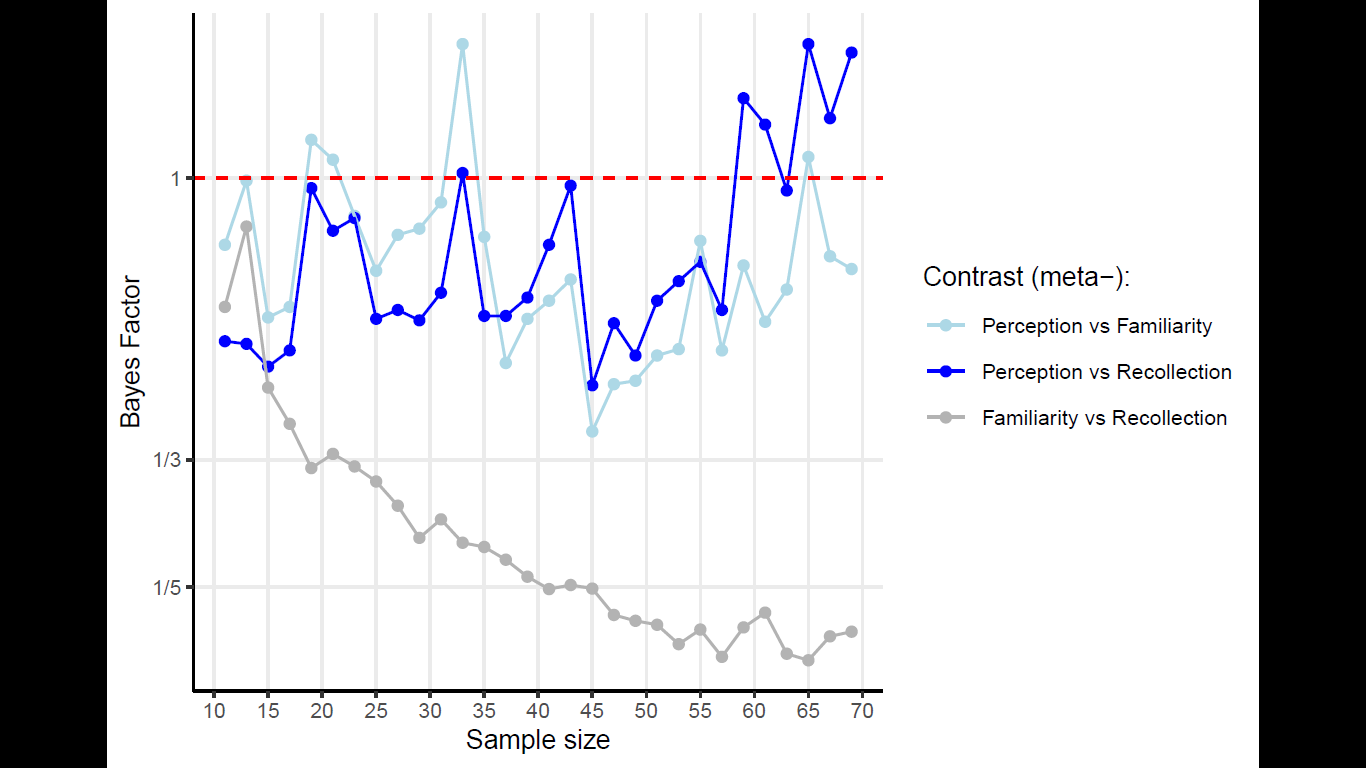

Figure S1. Bayesian sequential analysis of the interaction between task and group on metaperformance for each contrast: Visual detection vs Familiarity in blue, Visual detection vs Recollection in light blue, Familiarity vs Recollection in gray. Bayes Factors < ⅓ for the contrast between meta-familiarity and meta-recollection are evidence for an absence of a specific deficit in memory tasks.

##### Exclusions

Visual inspection led to participant exclusion in the following situations: 1) a pattern of first-order performance resulting from a strong response bias, revealed by high (resp. low) and stable first-order performance across all tasks for all non-null stimulus strength, and low (resp. high) performance across all tasks in catch trials (i.e. null evidence), strongly suggesting that those participants did not do the task properly; 2) no variability in confidence judgments (leading to the impossibility to compute indices of metacognitive performance). Accordingly, we excluded 3 patients with extreme values of criterion as well as weak sensitivities across all tasks; 3 control participants with a ceiling effect on confidence ratings (i.e. no variance in their responses); one patient due to the misuse of the confidence scale, revealed by a bimodal distribution of confidence ratings.

#### Recruitment procedure

Patients were recruited from community mental health centers and outpatient clinics in Versailles and Grenoble and were included if they met the criteria for a diagnosis of schizophrenia or schizoaffective disorders according to the DSM-5 during a diagnostic interview. Healthy volunteers between 18 and 65 years old were recruited from the general population, matched to the patients for age, gender, and education. All participants had normal or corrected-to-normal vision. Exclusion criteria for both groups comprised an estimated IQ (from Wechsler Adult Intelligence Scale IV matrix subtest^1^) strictly lower than 2 standard deviations below the mean of the general population; substances or alcohol dependence within the past 6 months and current; or prior history of untreated significant medical illness or of neurological illness. The control group was screened for current psychiatric illness during a diagnostic interview and participants were excluded in case they met criteria for any mental disorders according to the DSM-V.

#### Clinical and neuropsychological evaluation

We used the French National Adult Reading Test^2^ to assess premorbid IQ, and the matrix reasoning subtest from the WAIS-IV to exclude participants scoring lower than two standard deviations below the mean of the general population.

#### Stimuli

Using a 2-D Fourier Transform, the phase of each face image was randomized to create random noise backgrounds with spatial frequencies identically distributed. This noise background was grayscaled (familiarity and visual detection task) or presented in blue or red to provide contextual information (recollection task). Luminosity was balanced between blue and red noise backgrounds using the following formula: L = 0.30*R + 0.59*G + 0.11*B. (where R, G and B stand for the blue, green and red channels).

#### Randomization

Within each trial, the sequence of faces during the encoding phase was pseudo-randomized regarding gender and background color to get 2 male faces and 2 women faces (hence 6 combinations possible), 2 blue and 2 red backgrounds (6 combinations), totalizing 36 possible combinations. Participants were asked to provide confidence judgments only in session 2.

#### Structure of the experiment

The rationale behind the split into two sessions is the following: a pilot study had previously demonstrated that individual performances were similar between familiarity and recollection tasks. Thus, the first session of the experiment started with the assessment of familiarity and recollection performance for each lag. In order to match perceptual performance on memory performance, we determined a visual psychometric curve from which we selected perceptual intensities corresponding to the memory performances obtained previously (see Figure S2 D.).

The first session had three parts, in the following order: 1) 5 blocks of 40 memory trials (familiarity and recollection condition randomly interleaved), 2) 35 trials in the visual detection task with a 1up/2down staircase procedure to determine the 71% detection threshold, and 3) 3 blocks of 80 trials in the visual detection condition, with 10 contrast levels relative to the detection threshold (relative intensities: [0, 0.7, 0.8, 0.9, 1.0, 1.1, 1.2, 1.3, 1.4, 1.5]). A psychometric curve was fitted for each participant (see Figure S2 A., B, C.).

To match perceptual performance with memory performance, we used the psychometric curve obtained from the end of session 1 as follows: for each perceptual trial in session 2, we selected the stimulus intensities corresponding numerically to the minimum and maximum of memory performance. Adding two equidistant performance levels between the minimum and maximum, we obtained four contrast levels to map with the four memory stimulus strengths (or lags), in terms of resulting performance (see Figure S2 D.).

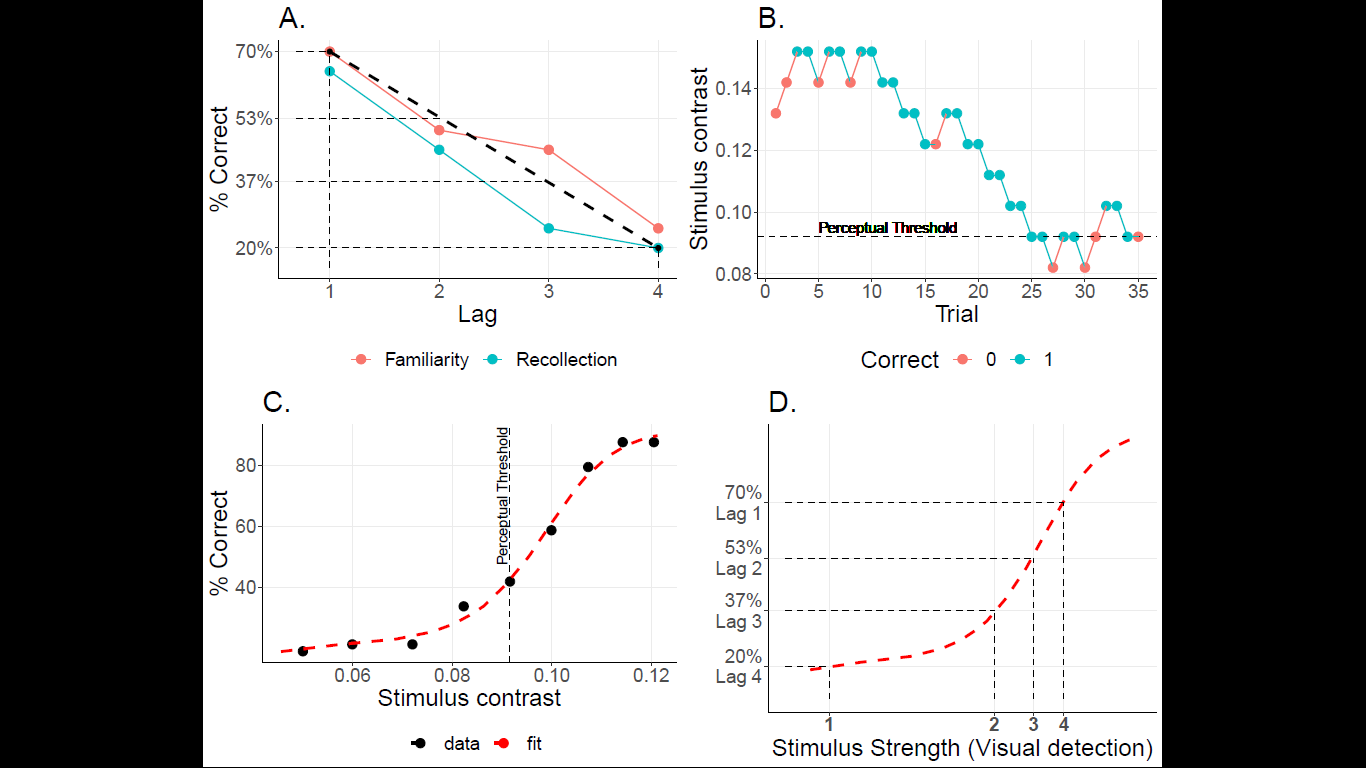

Figure S2. Experimental procedure for performance-matching. A. Memory performance obtained from one participant, during session 1. Memory performances to be matched by perceptual performance were computed at this stage for each stimulus strength. To ensure a wide range of performance, we took the minimum performance at Lag 4, the maximum performance at Lag 1, and equidistant values for Lag 2 and Lag 3. B: Visual detection staircase from one participant, during session 1. C: Psychometric curve obtained from one participant, during session 1. D. Illustration of the performance-matching procedure for one subject. Perceptual intensities were determined from memory performance measured in session 1 projected onto the individual psychometric curve, and updated online during session 2.

#### Bayesian analysis: prior specification

Priors were defined as follows:

We defined weakly informative Gaussian priors in the direction of a lower task performance for patients compared to controls (mean = -0.5, SD = 1); equivalent accuracy across the three tasks for patients (mean = 0, SD = 1) as well as for healthy controls (mean = 0, SD = 1); increased accuracy for higher stimulus strength (mean = 0.5, SD = 1); a positive regression slope of accuracy as a function of confidence for control participants, which served as baseline metacognitive performance (mean = 0.5, SD = 1), a metacognitive deficit among individuals with schizophrenia (lower regression slopes for patients compared to healthy controls: interaction confidence * group, mean = -0.1, SD = 1). Due to the absence of evidence for a specific metacognitive impairment among individuals with schizophrenia (studies controlling for first-order performance across multiple domains), the prior for the double interaction confidence * group * task was centered on 0 (mean = 0, SD = 1).

#### Response times

Response times were log-transformed and modeled with a bayesian linear mixed-effects regression, with accuracy (binary categorical variable: correct or incorrect), standardized confidence (continuous variable), group (binary categorical variable: controls vs patients), evidence (ordinal variable with 4 levels, i.e. a common scale for memory lag levels and perceptual contrast levels. Stimulus strength is symmetrical to difficulty), task (categorical variable: perception, familiarity, recollection) as fixed effects, and a full random effect structure.

Formula:

log(RT) ~ accuracy * confidence * group * task * evidence

+ (confidence*task*evidence | participant)	(2)

Based on Faivre and colleagues^3^, we expected to replicate the following effects on response times: longer response times for patients compared to healthy control participants, shorter response times for correct vs. incorrect responses, a negative correlation between response times and confidence, a lower link between response times and response correctness among individuals with schizophrenia compared to healthy controls, a lower link between response times and confidence ratings among individuals with schizophrenia compared to healthy controls. According to these predictions, we defined the following Gaussian priors: shorter response times for correct vs. incorrect responses (mean = -0.1, SD = 1), longer response times for patients compared to healthy control participants (mean = 0.1, SD = 1); similar response times across the three tasks for patients (mean = 0, SD = 1) as well as for healthy controls (mean = 0, SD = 1); shorter response times for higher stimulus strength (mean = -0.1, SD = 1); a negative correlation between response times and confidence ratings (mean = -0.1, SD = 1), a lower link between response times and accuracy among individuals with schizophrenia compared to healthy controls (mean = 0.1, SD = 1), and a lower link between response times and confidence ratings among individuals with schizophrenia compared to healthy controls (mean = 0.1, SD = 1).

#### Domain-generality of metacognition

To assess whether metacognitive performance correlated across tasks while avoiding spurious correlations due to group-level shrinkage in hierarchical models, independent generalized mixed models were conducted for each subject and each task (i.e. 3 models per subject) as follows:

accuracy ~ confidence * evidence	(3)

Under the assumption of a domain-general architecture of metacognition^4^, we expected pairwise task metaperformance correlations. According to the disconnection hypothesis in schizophrenia^5^, we expected lower pairwise task-metaperformance correlations among individuals with schizophrenia compared to healthy controls.

#### Correlation with clinical scores

Robust linear regressions were performed to explore the correlations between individual metacognitive performance (indicated by regression slopes between accuracy and confidence) and 1) demographic and neuropsychological scores (age, education level, premorbid IQ, depression (CDS), and WAIS matrix subtest standardized score), 2) clinical scores (only for patients: PANSS positive, negative and disorganization scores, cognitive insight (BCIS) and subjective cognitive functioning (SSTICS total and working memory scores).

### Results

#### First-order performance

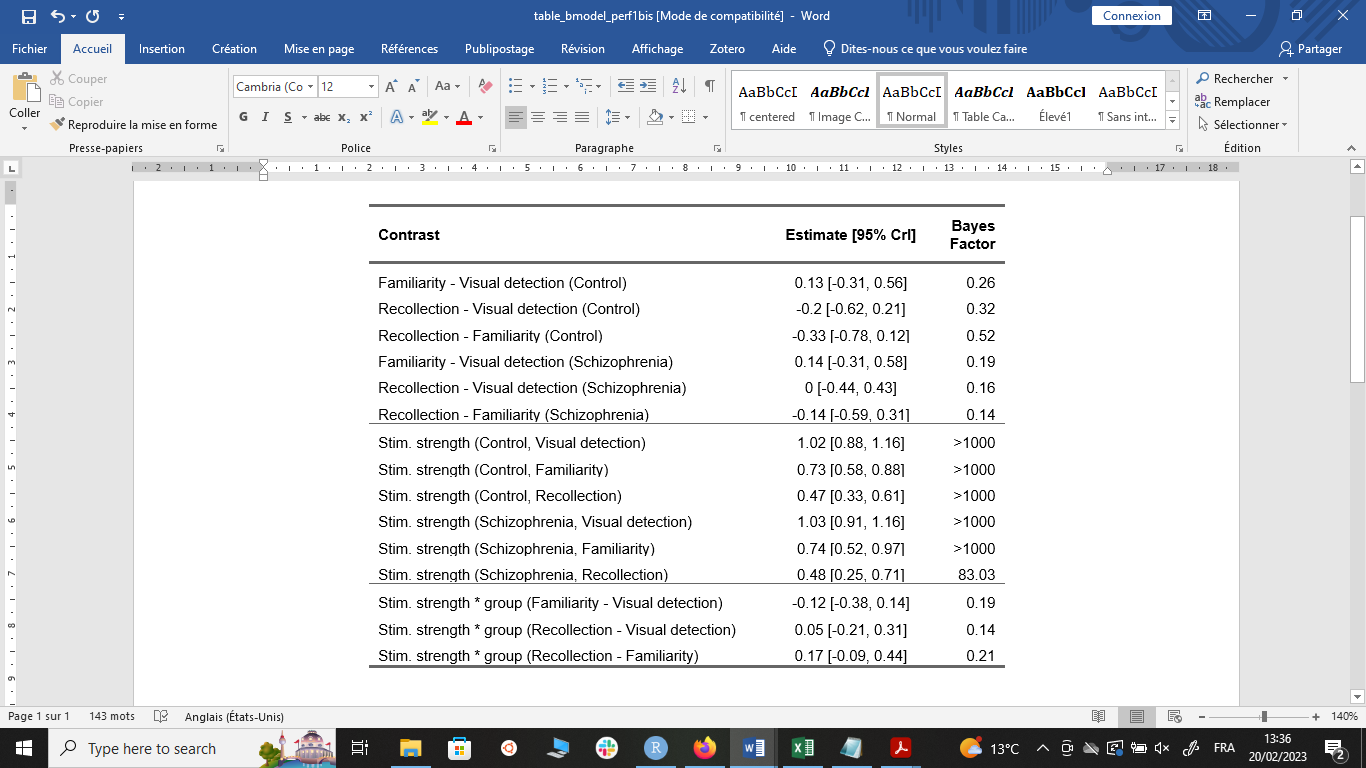

Table S1.

Plausible explanation of imperfect intra-individual performance matching:

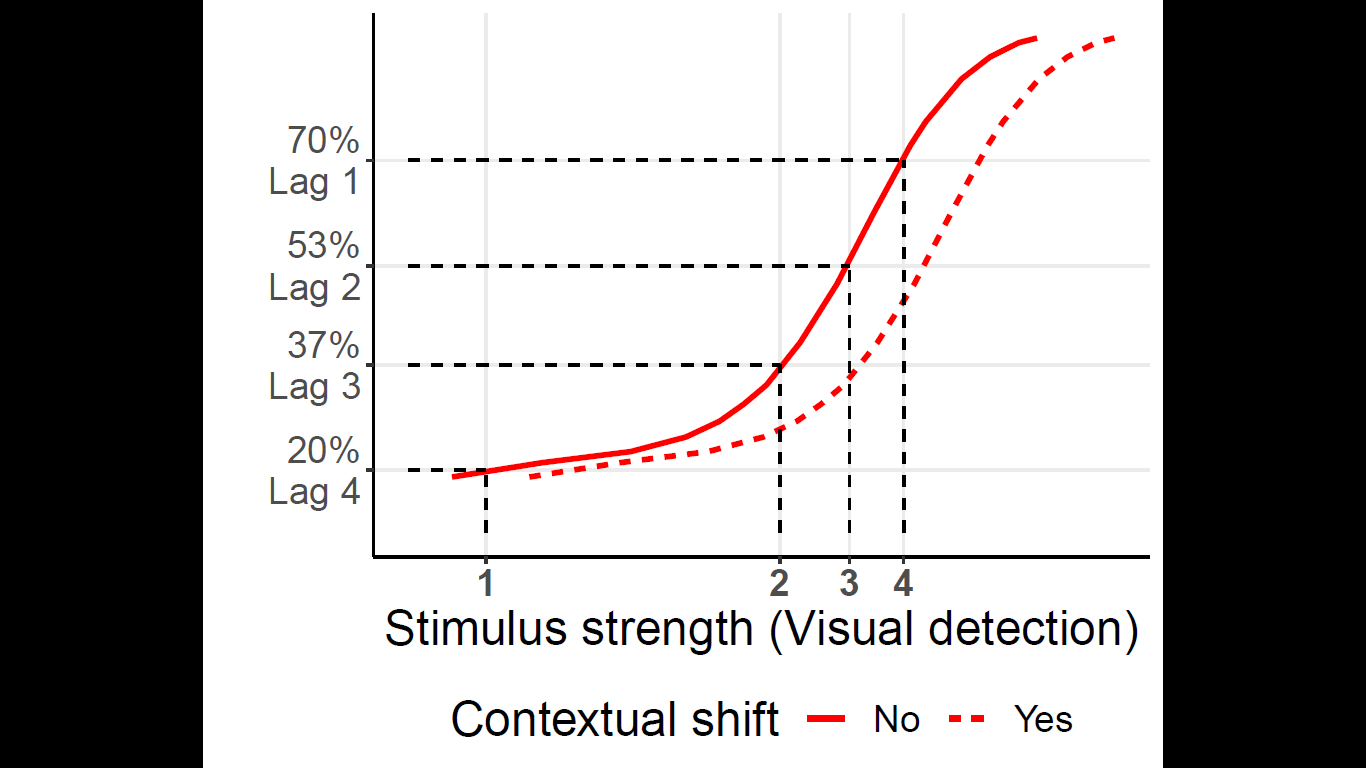

Figure S3. Reproduction of Figure S2 D., with an additional dashed psychometric curve rightward shifted compared to the solid curve, that would result from a contextual effect of high-contrast memory stimuli embedded within low-contrast visual detection stimuli during session 2.

#### Logistic regressions

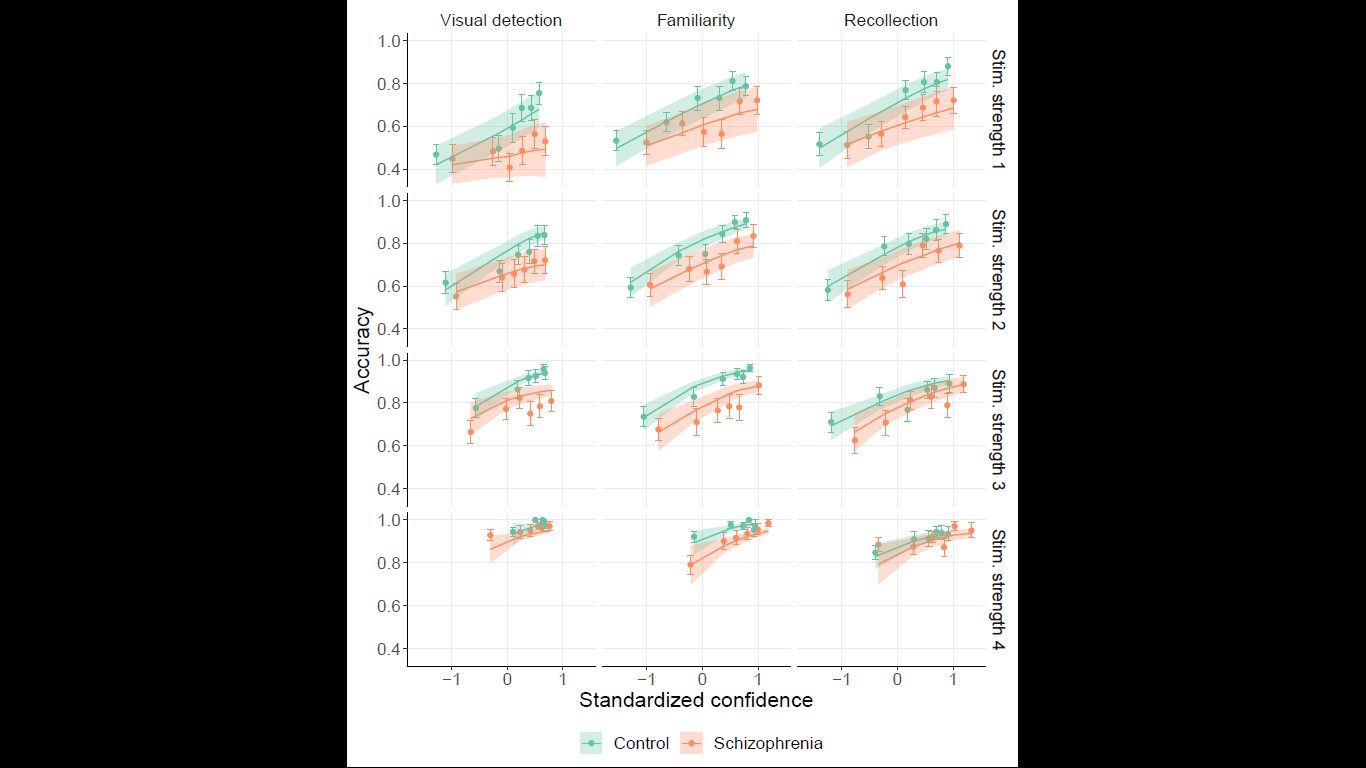

Figure S4: Accuracy as a function of standardized confidence, by group, task (columns), and stimulus strength (rows). Points and error bars indicate average accuracy and standard error of the mean, respectively, computed for each confidence bin. Solid lines and shaded areas represent model fit mean and 95% confidence interval, respectively.

#### Response times

On average, patients were slower to respond compared to healthy controls (0.19 [0.07, 0.31], BF = 257). Response times were shorter for correct responses compared to incorrect responses among individuals with schizophrenia (-0.08 [-0.10, -0.05], BF = 8000) and control participants (-0.23 [-0.26, -0.20], BF = 8000). However, correctness was less predictive of response times among patients compared to healthy controls, as indicated by the very strong evidence for the ‘correctness * group’ interaction (0.15 [0.11, 0.19], BF = 8000).

Response times were negatively correlated with confidence in both groups (patients: -0.19 [-0.24; -0.14], BF = 8000; controls: -0.21 [-0.24, -0.18], BF = 8000), meaning that participants were longer to respond when less confident, regardless of the group (as indicated by the weak and inconclusive ‘confidence * group’ interaction: 0.02 [-0.04, 0.07], BF = 2.44). However, the strength of the link between response times and confidence was higher among controls compared to individuals with schizophrenia for correct responses, but not for incorrect responses (Figure S5, ‘confidence * group * correctness’ double interaction: 0.08 [0.04, 0.13], BF = 614).

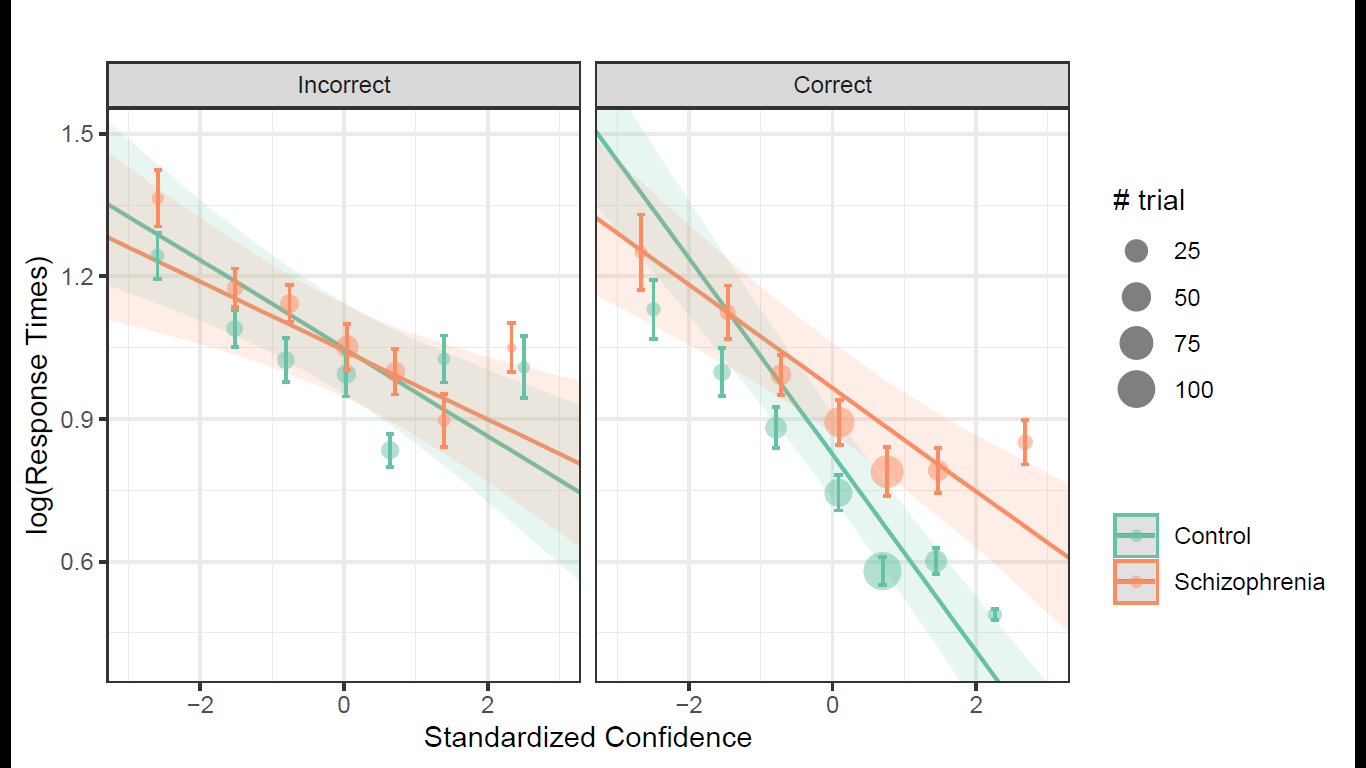

Figure S5: Response times (log-transformed) regressed on standardized confidence for incorrect responses (left panel) and correct responses (right panel). Points, size of points, and error bars indicate means, averaged number of individual trials, and standard errors, resp.; solid lines and shaded areas represent model fit means and 95% CrI, resp.

In both groups of participants, response times for correct responses were longer in the visual detection task compared to the familiarity task (Patients: -0.18 [-0.25, -0.12], BF = 8000, Controls: -0.16 [-0.21, -0.10], BF = 8000) and compared to the recollection task (Patients: -0.05 [-0.11, 0.02], BF = 6.71, Controls: -0.04 [-0.10, 0.02], BF = 6.71)(Figure S6).

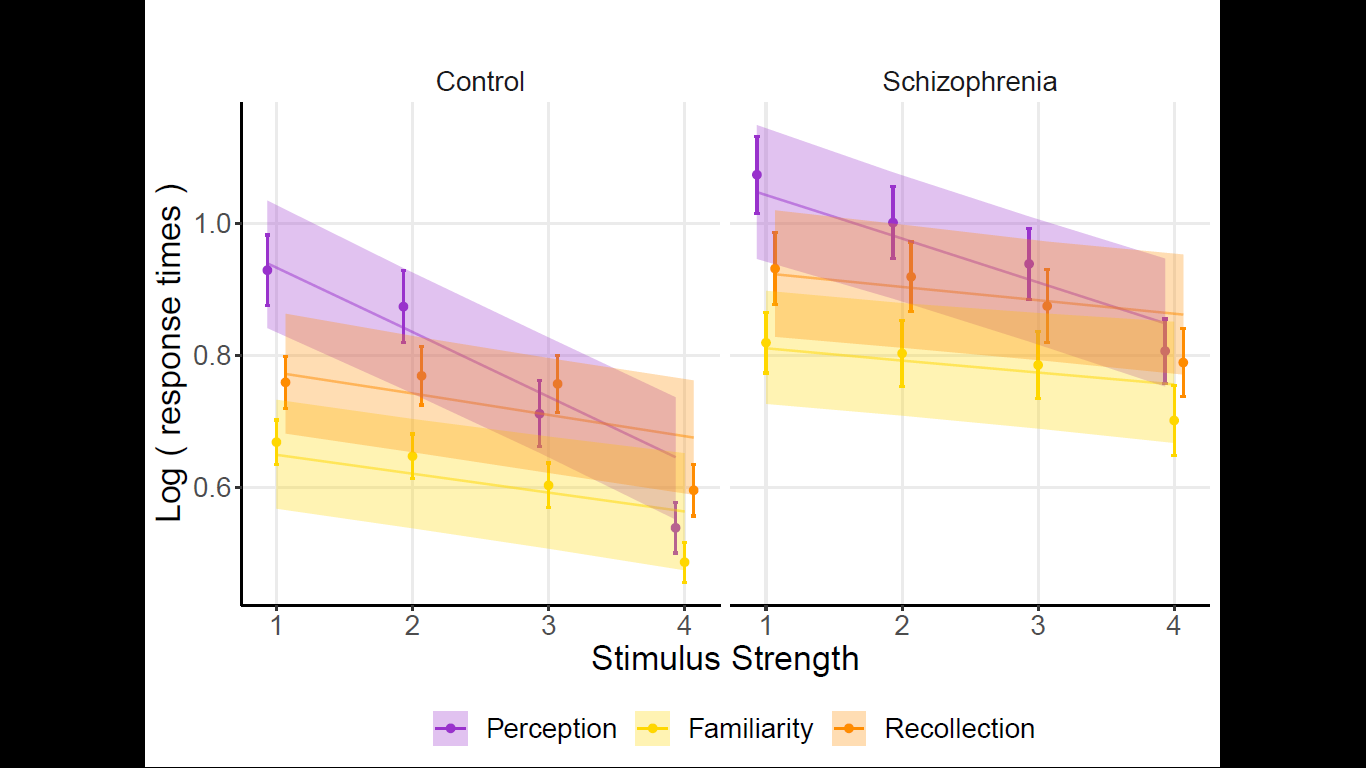

Figure S6: Log-Response times (only for correct responses) as a function of stimulus strength, for each tack and group. Points and error bars indicate average accuracy and standard error of the mean, respectively; solid lines and shaded areas represent model fit mean and 95% CrI, respectively.

#### Domain-generality

Interestingly, contrary to the notion that metacognition obeys domain-general rules, we found no pairwise correlations between indices of metacognitive sensitivity across tasks (Figure S7; Perception - Familiarity: estimate = -0.16, std err. = 0.37, statistic = -0.45, p = 0.65, BF = 0.36; Recollection - Familiarity: estimate = 0.08, std err. = 0.11, statistic = 0.69, p = 0.49, BF = 0.40; Recollection - Perception: estimate = -0.07, std err. = 0.27, statistic = -0.26, p = 0.79, BF = 0.33)).

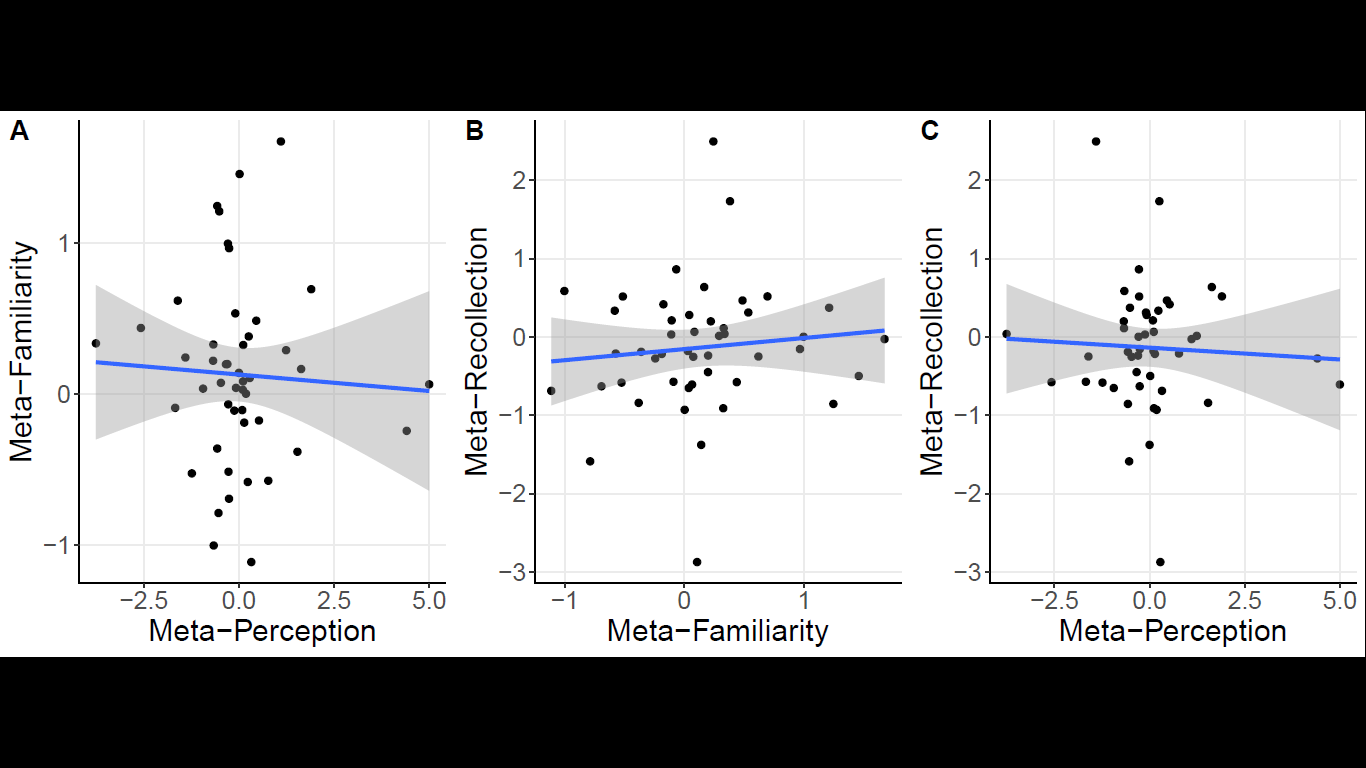

Figure S7: Pairwise task-metaperformance correlations. Dots represent individual regressions slopes (accuracy regressed on confidence). Solid lines and shaded areas represent the correlational fits, and standard errors, resp.

#### Correlation with clinical scores

Among patients, metacognitive performance tended to be positively correlated with cognitive insight scores (estimate = 0.03, std err. = 0.02, t = 1.75, p = 0.08, BF = 1.08), and was negatively correlated - yet with inconclusive evidence - with PANSS disorganized symptoms (estimate = -0.05, std err. = 0.02, t = -2.43, p < 0.05, BF = 1.09) and with PANSS negative symptoms (estimate = -0.04, std err. = 0.02, t = -2.23, p < 0.05, BF = 0.72). Other clinical scores - PANSS positive symptoms and subjective cognitive functioning (SSTICS) - were not correlated with metacognitive performance (Figure S8).

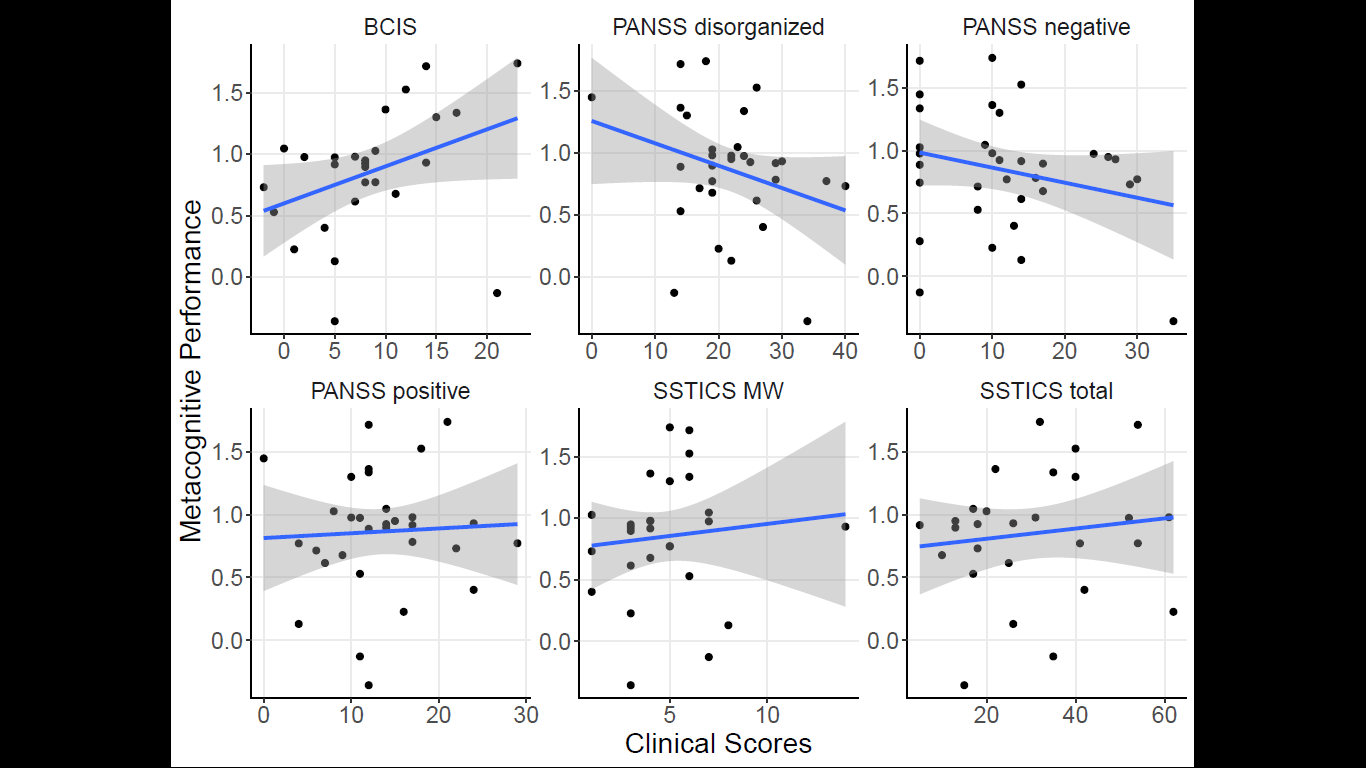

Figure S8: Metacognitive scores regressed on clinical scores among patients. Dots are individual regression slopes averaged across the three tasks. BCIS: Beck Cognitive Insight Scale; PANSS: Positive And Negative Symptoms in Schizophrenia, SSTICS WM: Subjective Scale To Investigate Cognition in Schizophrenia: Working Memory score.

Among demographic and neuropsychological variables, only the WAIS matrix subtest scores were correlated with metacognitive performance (Figure S9, estimate = 0.09, std err. = 0.03, t = 3.13, p < 0.01, BF = 415).

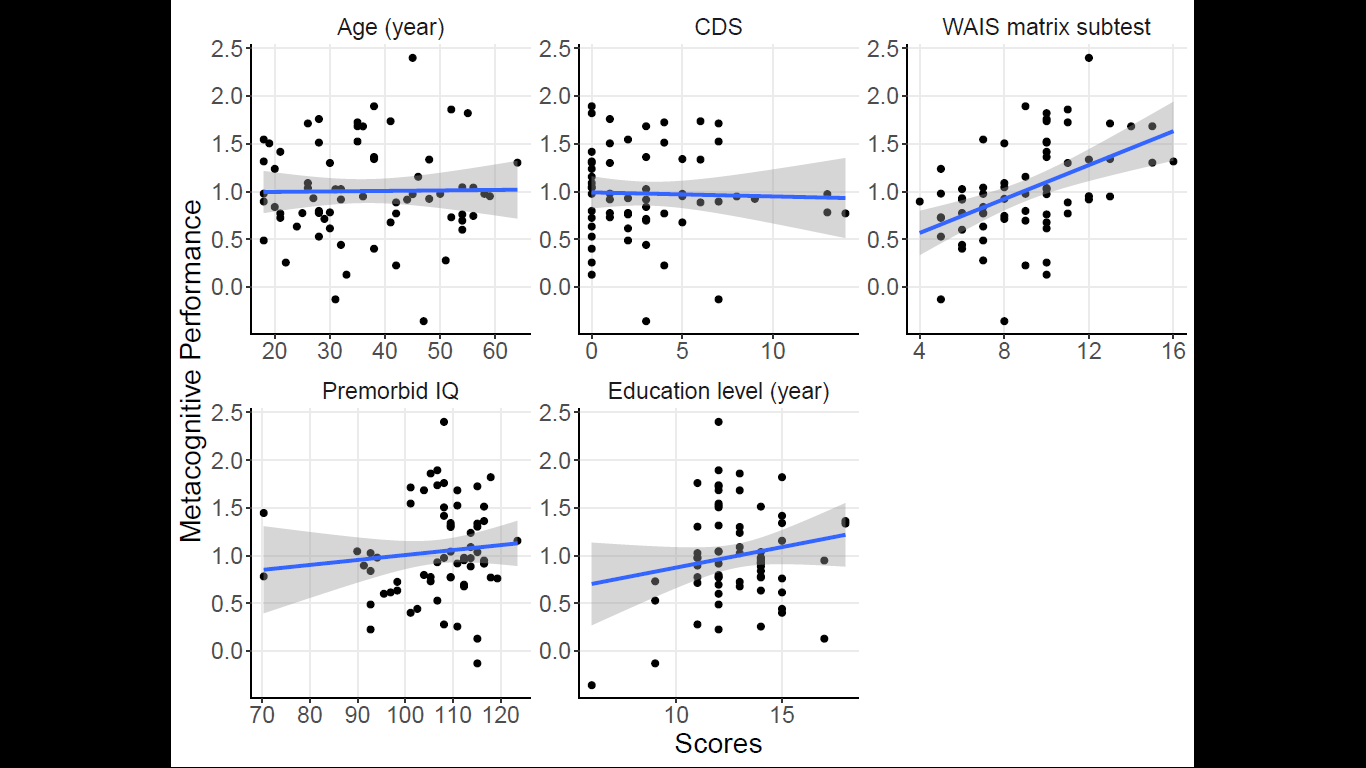

Figure S9: Metacognitive scores regressed on demographic data and neuropsychological scores (all participants). Dots are individual regression slopes averaged across the three tasks.

### References

1.	Wechsler D. Wechsler Adult Intelligence Scale--Fourth Edition. November 2012. doi:10.1037/t15169-000

2.	Mackinnon A, Mulligan R. Estimation de l’intelligence prémorbide chez les francophones. *L’Encéphale*. 2005;31(1):31-43. doi:10.1016/S0013-7006(05)82370-X

3.	Faivre N, Roger M, Pereira M, et al. Confidence in visual motion discrimination is preserved in individuals with schizophrenia. *J Psychiatry Neurosci*. 2021;46(1):E65-E73. doi:10.1503/jpn.200022

4.	Mazancieux A, Fleming SM, Souchay C, Moulin CJA. Is there a G factor for metacognition? Correlations in retrospective metacognitive sensitivity across tasks. *J Exp Psychol Gen*. 2020;149(9):1788-1799. doi:10.1037/xge0000746

5.	Friston KJ. The disconnection hypothesis. *Schizophr Res*. 1998;30(2):115-125. doi:10.1016/S0920-9964(97)00140-0
